# Postoperative Blood Pressure and Outcomes in Aneurysmal Subarachnoid Hemorrhage

**DOI:** 10.1101/2025.09.25.25336694

**Authors:** Ting Liu, Zhongxiao Wang, Xue Xia, Shengqi Hu, Shuang Wang, Wenqiang Li, Xinjian Yang

## Abstract

**Background:** Optimal postoperative blood pressure (BP) management remains unclear for hypertensive patients with aneurysmal subarachnoid hemorrhage (aSAH). We investigated associations of early postoperative BP levels, variability, and trajectories with 6 month outcomes.

**Methods:** Consecutive hypertensive patients after aSAH surgery were retrospectively analyzed. BP was measured four times daily for 3 days. Profiles included minimum systolic BP (SBP), mean arterial pressure, variability, and SBP trajectories. Minimum SBP was dichotomized at 140 mmHg to examine threshold effects. Multivariable logistic regression assessed associations between BP metrics and functional outcome. Causal mediation analysis evaluated the indirect effect of delayed cerebral ischemia (DCI) on the association between minimum SBP (140 mmHg cutoff) and outcome.

**Results:** Among 702 patients, lower SBP in the first 3 days independently predicted poor outcomes (OR = 0.98). Maintaining SBP ≥140 mmHg was linked to lower risks of unfavorable outcomes (OR = 0.44) and DCI (OR = 0.49). Mediation analysis indicated 35% of this effect was explained by reduced DCI. Higher BP variability, measured by standard deviation and coefficient of variation, correlated with poor outcomes. Trajectory analysis showed that moderately rising SBP (≥140 mmHg) was associated with the favorable outcomes.

**Conclusions:** Maintaining SBP ≥140 mmHg with stable variability in the early postoperative period was associated with favorable functional outcomes, partly through reducing DCI. These findings highlight the importance of adequately elevated and stable BP management after aSAH surgery.

## INTRODUCTION

Aneurysmal subarachnoid hemorrhage (aSAH) is a subtype of hemorrhagic stroke, with an estimated 697,000 new cases and 353,000 deaths worldwide in 2021, imposing a substantial burden on patients and healthcare systems1–3. Despite advances in endovascular therapy and intensive care, the mortality rate remains 25– 30%, and many survivors are left with long-term neurological deficits4.

Blood pressure (BP) elevation and fluctuations are common after aSAH, and both absolute BP levels and variability are strongly associated with rebleeding, delayed cerebral ischemia (DCI), and poor functional outcomes5–8. BP management is therefore crucial, particularly in patients with aSAH who present with elevated BP. However, management in this population is complex because uncontrolled hypertension increases the risk of rebleeding, whereas excessive BP reduction may compromise cerebral perfusion and trigger DCI9. Before securing the aneurysm, current consensus recommends keeping systolic BP below 160-180 mmHg to reduce the risk of re-rupture10–12. In contrast, evidence on postoperative BP parameters and their association with outcomes is limited, and optimal management strategies after aneurysm repair remain controversial. Triple-H therapy (hypertension, hypervolemia, hemodilution) is no longer recommended because of potential harm, including neurogenic cardiac injury, posterior reversible encephalopathy syndrome, and seizures1. Furthermore, the prematurely terminated HIMALAIA trial showed that induced hypertension did not improve long-term outcomes and was associated with a higher incidence of serious adverse events13. A small retrospective studies have also suggested that while BP lowering may be reasonable in patients with hypertension, it should be avoided during the vasospasm period to preserve cerebral perfusion14.

Against this background, blood pressure profiles during the first three postoperative days in aSAH patients with elevated BP were analyzed to characterize BP levels, variability, and trajectory patterns, and to assess their associations with 6-month functional outcomes.

## METHODS

### Patient population

Patients were included if they met the following criteria: (1) subarachnoid hemorrhage confirmed by computed tomography; (2) an intracranial aneurysm identified by computed tomography angiography and digital subtraction angiography as the source of bleeding; (3) admission within 24 hours of symptom onset and treatment with surgical clipping or endovascular embolization within 72 hours after admission; and (4)having at least three blood pressure recordings per day throughout the observation period. Exclusion criteria were severe systemic comorbidities (such as significant hepatic, renal, or cardiac dysfunction or hematologic disorders), incomplete clinical, imaging, or follow-up data, and postoperative SBP < 140 mmHg. A flowchart of the patient selection process is shown in **Figure 1**. This study was conducted at XX Hospital, between January 2016 and December 2022, with ethical approval (KY 2023-261-01). Informed consent was obtained from all participants or their legally authorized representatives.

**Figure 1:**
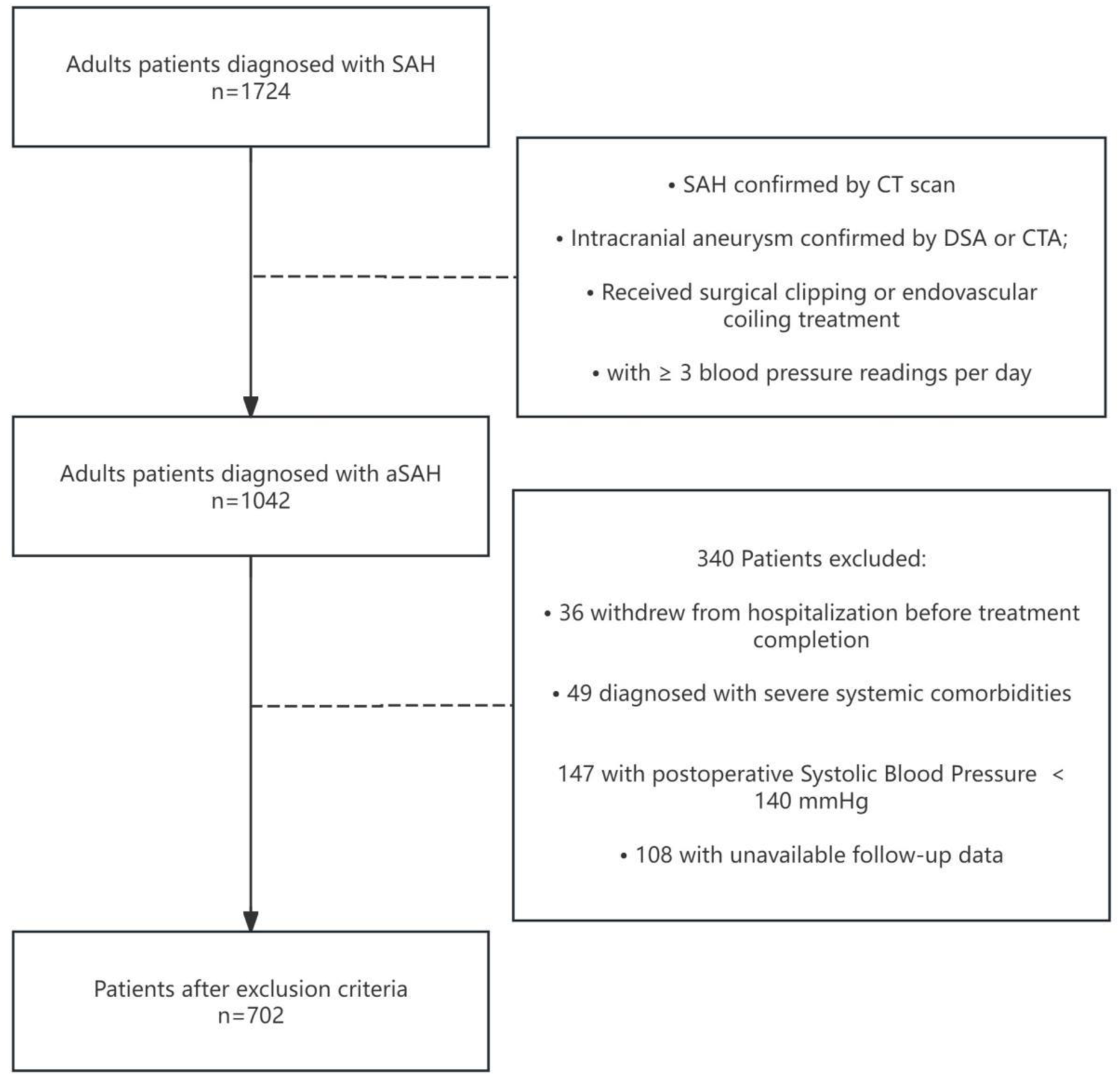
Flow diagram of study patients. SAH, subarachnoid hemorrhage; aSAH, aneurysmal subarachnoid hemorrhage; CT, computed tomography; CTA, computed tomographic angiography; DSA, digital subtraction angiography.

### Patient management and treatment

All patients received care according to national and international guidelines 10–12. After initial assessment in the emergency department, they were admitted to either a specialized neurovascular ward or the intensive care unit based on clinical severity. For aSAH cases, treatment strategy, microsurgical clipping or endovascular therapy, was determined through multidisciplinary discussion between vascular neurosurgeons and interventional specialists. When endovascular therapy was selected, simple coiling without stent assistance was preferred; if stent use was necessary, antiplatelet therapy was administered intraoperatively and continued after the procedure. In patients with a large intracranial hematoma, hematoma evacuation was performed concurrently with aneurysm clipping to reduce mortality and improve neurological outcomes. Patients with increased intracranial pressure or suspected hydrocephalus were managed with an external ventricular drain or lumbar drainage. All patients were managed in the intensive care unit with standard vasospasm prophylaxis, including nimodipine, volume optimization, and, when indicated, induced hypertension15.

### Data collection and definitions

Patient data were extracted from electronic medical records, including demographic information (age, sex, smoking and alcohol history), comorbidities (hypertension, hyperlipidemia, diabetes mellitus), Hunt–Hess grade (HH grade), modified Fisher grade (mFisher grade), aneurysm location, Treatment modalities (clipping and coiling, use of antihypertensive medication. Blood pressure was recorded four times daily during the first 3 postoperative days. For each patient, the lowest systolic blood pressure (SBP) and mean arterial pressure (MAP) were extracted for the intervals 0-24 h, 24-48 h, 48-72 h, as well as cumulatively for 0-48 h and 0-72 h after surgery. These minimum SBP and MAP values, considered proxies for cerebral perfusion, were analyzed as primary variables to assess their associations with 6-month functional outcomes16, 17.

### Definition of blood pressure variability Indices

Blood pressure variability (BPV) was evaluated from SBP measurements within the first 72 hours after surgery using four validated indices8, 18, 19: (1) standard deviation (SD), representing the overall dispersion of SBP values; (2) average real variability (ARV), calculated as the average of the absolute differences between consecutive SBP readings, thereby capturing the magnitude of short-term fluctuations while accounting for their temporal order, (3) successive variation (SV), reflecting the variability between consecutive SBP measurements, and (4) coefficient of variation (CV), defined as SD divided by the mean SBP and expressed as a percentage, which normalizes dispersion to the mean. BPV was firstly analyzed as a continuous variable. For additional analyses, Patients were categorized into three groups as low, moderate, and high BPV groups by tertiles20.

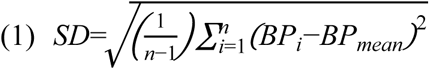

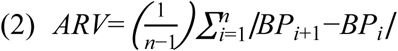

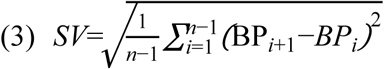

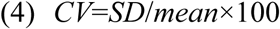

### Identification of blood pressure trajectories

Group-based trajectory modelling was applied to identify distinct postoperative SBP patterns during the first 3 days after surgery21. This method, a specialized form of finite mixture modeling, groups individuals who follow similar temporal SBP trajectories. Models specifying one to five classes and incorporating different polynomial functions were fitted. The optimal number of trajectories was determined primarily using the Bayesian Information Criterion (BIC), while ensuring that each class contained at least 5% of the study population and achieved a mean posterior probability greater than 0.7022. Each patient was then assigned to the trajectory class for which they had the highest posterior probability.

### Outcome Assessment

The primary outcome was favorable functional outcome at 6-months post-discharge, defined as an mRS score of 0 to 2. The secondary outcome was the occurrence of DCI during hospitalization, defined in accordance with the criteria used by Vergouwen et al23.

### Statistical analysis

Continuous variables were reported as mean±standard deviation or median (interquartile range), and categorical variables as counts and percentages (n[%]). The Shapiro-Wilk test was used to assess normality. Continuous variables were compared using the Student’s t-test for normally distributed data or the Mann-Whitney U-test for non-normally distributed data. Categorical variables were compared using the χ² test or Fisher’s exact test. Variables with P < 0.10 in univariate analyses were entered into a multivariable logistic regression model to identify independent predictors of functional outcome and DCI, using a stepwise backward elimination procedure for model selection. Causal mediation analysis was performed to evaluate whether DCI mediates the association between postoperative SBP < 140 mmHg and unfavorable outcome24. Statistical analyses were conducted using SPSS Statistics 27 (IBM, Armonk, NY) and R 4.2.0 software. All tests were two-sided, with statistical significance defined as p<0.05.

## RESULTS

### Patients’ characteristics

A total of 702(82.7%) consecutive patients with postoperative hypertension were enrolled in the study, while 147 patients (17.3%) without postoperative hypertension were excluded **(Figure 1)**. 501 patients (71.37%) received antihypertensive therapy after surgery. Baseline characteristics are outlined in **Table 1**. At the 6 month follow up, 169 patients (24.1%) had an unfavorable outcome. Patients with an unfavorable outcome were older and more likely to have a Hunt-Hess grade >3 and a modified Fisher grade >2 at admission. Surgical clipping and antihypertensive therapy were also more frequent in this group compared with those with a favorable outcome (all P < 0.05). During the first 72 hours after surgery, 12 consecutive SBP measurements had median values ranging from approximately 135 to 145 mmHg, and MAP measurements ranged from approximately 95 to 105 mmHg across time points **(Table S1-2 and Figure S1-2)**.

**Table 1.**
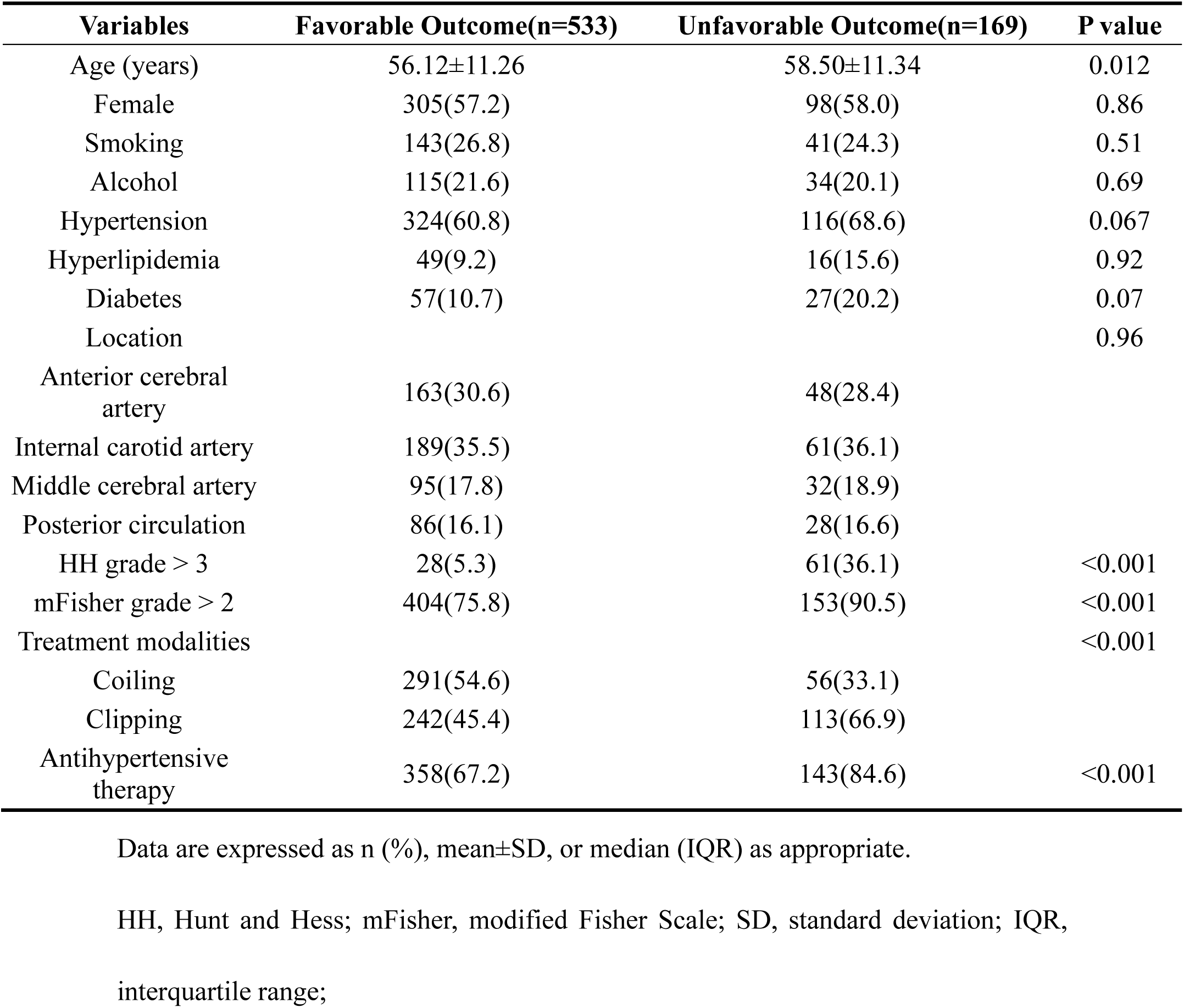
Baseline characteristics and outcomes of 702 patients with aneurysmal subarachnoid hemorrhage.

| Variables | Favorable Outcome(n=533) | Unfavorable Outcome(n=169) | P value |
| --- | --- | --- | --- |
| Age (years) | 56.12±11.26 | 58.50±11.34 | 0.012 |
| Female | 305(57.2) | 98(58.0) | 0.86 |
| Smoking | 143(26.8) | 41(24.3) | 0.51 |
| Alcohol | 115(21.6) | 34(20.1) | 0.69 |
| Hypertension | 324(60.8) | 116(68.6) | 0.067 |
| Hyperlipidemia | 49(9.2) | 16(15.6) | 0.92 |
| Diabetes | 57(10.7) | 27(20.2) | 0.07 |
| Location |  |  | 0.96 |
| Anterior cerebral artery | 163(30.6) | 48(28.4) |  |
| Internal carotid artery | 189(35.5) | 61(36.1) |  |
| Middle cerebral artery | 95(17.8) | 32(18.9) |  |
| Posterior circulation | 86(16.1) | 28(16.6) |  |
| HH grade > 3 | 28(5.3) | 61(36.1) | <0.001 |
| mFisher grade > 2 | 404(75.8) | 153(90.5) | <0.001 |
| Treatment modalities |  |  | <0.001 |
| Coiling | 291(54.6) | 56(33.1) |  |
| Clipping | 242(45.4) | 113(66.9) |  |
| Antihypertensive therapy | 358(67.2) | 143(84.6) | <0.001 |

Data are expressed as n (%), mean±SD, or median (IQR) as appropriate.
HH, Hunt and Hess; mFisher, modified Fisher Scale; SD, standard deviation; IQR,
interquartile range;

### Association Between Early Postoperative Minimum SBP and Functional Outcomes, With Mediation by DCI

Correlation analysis demonstrated a strong association between SBP and MAP across all time points **(Figure S3)**. Given this correlation and the greater clinical relevance and practicality of SBP, SBP was used as the primary variable, while MAP-related analyses are presented in **Figure S4**. During the first 3 postoperative days, the daily minimum SBP were analyzed. On postoperative day 3, patients with an unfavorable outcome exhibited significantly lower minimum SBP compared with those with a favorable outcome (P = 0.004). Interestingly, SBP remained relatively stable over time in the favorable outcome group, whereas the unfavorable outcome group showed a progressive decline across days 1-3 **(Figure 2A)**. Similarly, in univariable analysis, lower minimum SBP during the first 3 postoperative days was associated with unfavorable outcome, and this association remained significant after multivariable adjustment (SBP: OR = 0.97, 95% CI 0.96-0.99, P < 0.001; **Figure 2B-C**).

**Figure 2:**
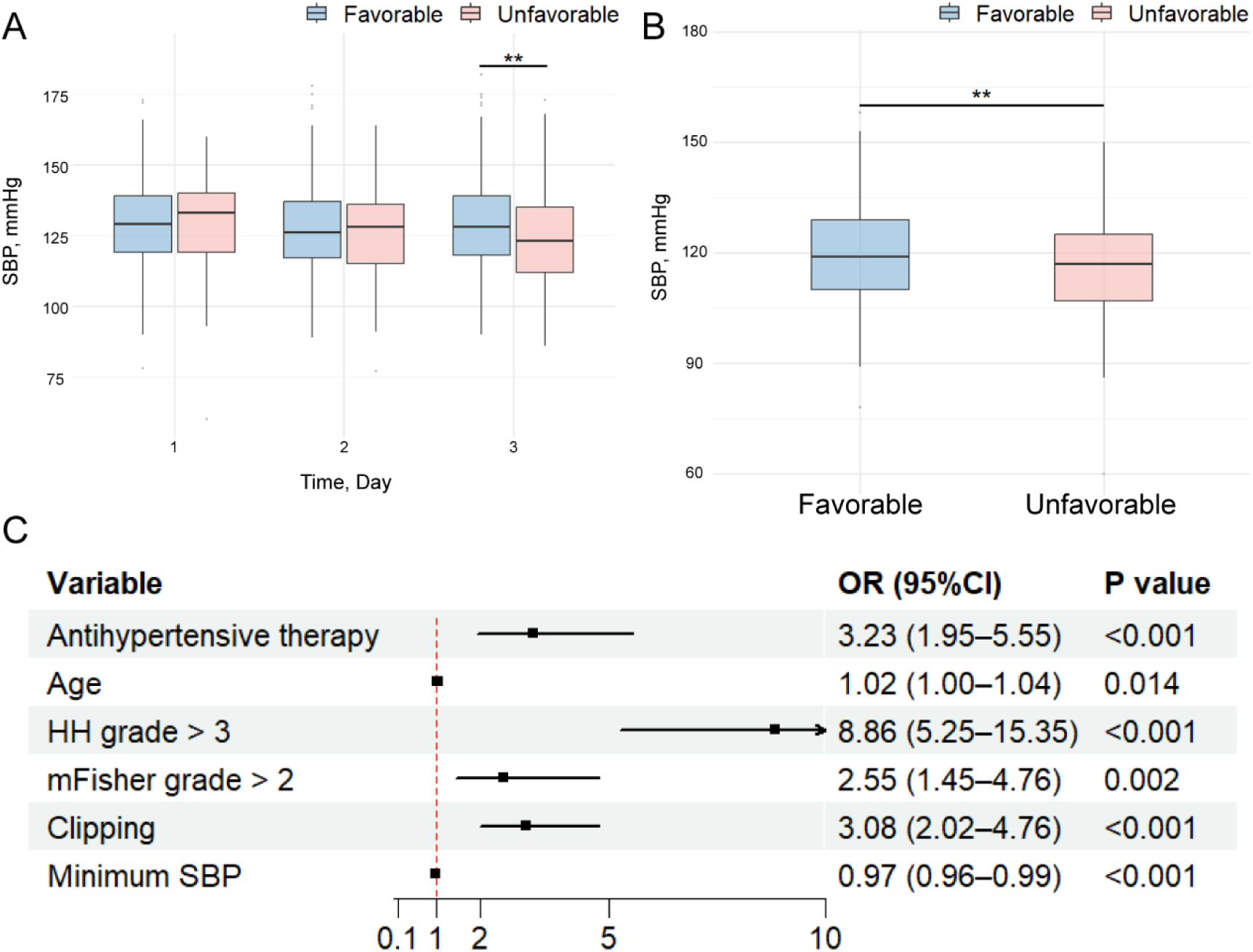
Association between postoperative systolic blood pressure (SBP) and clinical outcomes in patients with aneurysmal subarachnoid hemorrhage (aSAH). (A) Boxplots of SBP over the first three postoperative days stratified by favorable and unfavorable outcomes. (B) Boxplots of minimum SBP within the first 3 day stratified by outcomes. (C) Forest plot of multivariable logistic regression analysis showing independent predictors of unfavorable outcome. * P < 0.05; ** P < 0.01

To further explore prognostic thresholds, the lowest SBP within three days was dichotomized at 140 mmHg. Patients with a minimum SBP ≥ 140 mmHg had a lower rate of unfavorable outcomes compared with those with a minimum SBP < 140 mmHg (14.29% vs. 25.16%, P=0.05; **Figure 3A**). This association remained significant in multivariable logistic regression (OR = 0.44, 95% CI 0.19–0.92, P = 0.04; **Figure 3B**). Similarly, patients with a minimum SBP ≥ 140 mmHg also had a significantly lower incidence of DCI compared with those with SBP < 140 mmHg (OR = 0.49, 95% CI 0.24-0.95, P = 0.04; **Figure 3C-D**). Causal mediation analysis indicated that maintaining SBP ≥ 140 mmHg was associated with a −9.97% change in poor outcome (95% CI −18.31 to −3.06; P = 0.01), with −3.49% (95% CI −7.15 to −0.62; P = 0.03) mediated through DCI, accounting for 35.0% of the total effect (95% CI 8.12 to 69.92; P = 0.04), and −6.48% (95% CI −13.10 to −1.14; P = 0.04) representing a direct effect independent of DCI **(Figure 3E-F)**.

**Figure 3:**
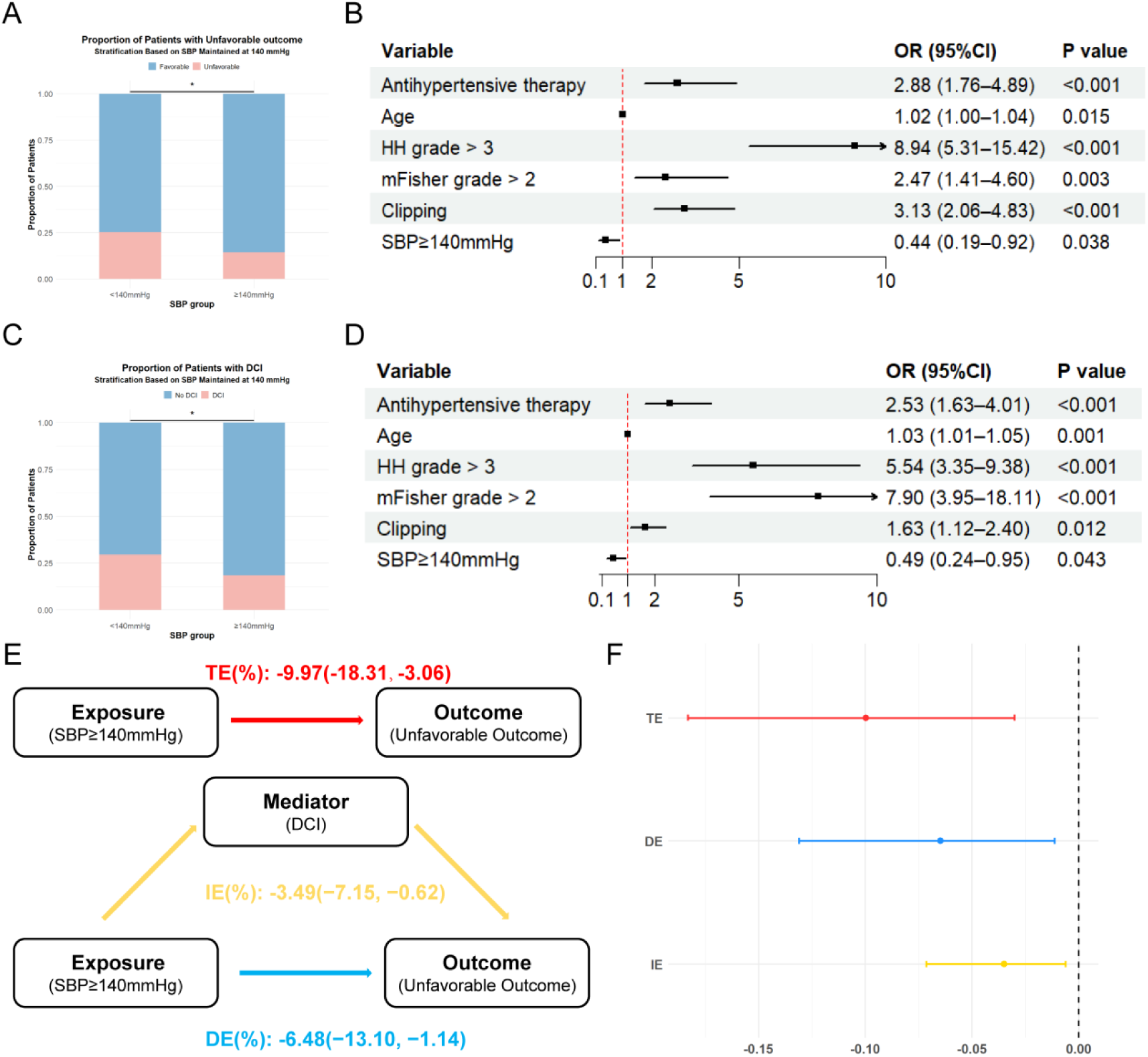
Association between postoperative systolic blood pressure (SBP) and outcomes, with delayed cerebral ischemia (DCI) as a mediator, in aneurysmal subarachnoid hemorrhage (aSAH). (A) Proportion of patients with unfavorable outcomes stratified by minimum SBP < 140 mmHg versus ≥ 140 mmHg within the first 72 h postoperatively. (B) Forest plot of multivariable logistic regression analysis showing independent predictors of unfavorable outcome. (C) Proportion of patients developing DCI stratified by minimum SBP < 140 mmHg versus ≥ 140 mmHg. (D) Forest plot of multivariable logistic regression analysis showing independent predictors of DCI. (E) Conceptual diagram of causal mediation analysis assessing the total effect (TE), direct effect (DE), and indirect effect (IE) of SBP ≥ 140 mmHg on unfavorable outcome through DCI. (F) Effect estimates and 95% confidence intervals for TE, DE, and IE from the mediation model. *P < 0.05.

### Association Between Postoperative BP Variability and Functional Outcomes

The association between postoperative BPV and functional outcomes was further examined using four BPV indices (SD, CV, ARV, and SV) categorized into tertiles(**Table S3**). In univariable analysis, higher SD and CV were associated with an increased risk of unfavorable outcomes, whereas ARV and SV showed no significant associations. These associations for SD (OR = 1.82, 95% CI 1.10-3.01; P = 0.02) and CV (OR = 2.17, 95% CI 1.34-3.52; P = 0.002) persisted in multivariable analysis(**Table S4**).

### Association Between Postoperative SBP Trajectories and Functional Outcomes

Trajectory models with one to five classes were constructed and compared. The five-class model was selected based on the lowest BIC, high mean posterior probabilities (>0.7 for all classes), and adequate class sizes (each >5% of the classes). The SBP trajectory patterns for models with 2 to 4 classes are shown in **Figure S5A-C**. In the final 5-class model, five distinct trajectories were identified: Group 1, moderate SBP with a gradual increase; Group 2, moderate SBP with a gradual decrease; Group 3, persistently high SBP; Group 4, moderately high SBP with a gradual decline; and Group 5, low SBP with a further decline **(Figure S5D)**. Multivariable analysis showed that, using Group 1 as the reference, the risk of unfavorable outcomes was higher in Group 2 (OR 2.27, 95% CI 1.17–4.39; P = 0.02), Group 4 (OR 2.37, 95% CI 1.16–4.86; P = 0.02), and Group 5 (OR 2.48, 95% CI 1.23–4.99; P = 0.01), while Group 3 did not reach statistical significance **(Table S5)**. Overall, trajectories characterized by a decline in SBP were associated with worse outcomes, while a moderate and gradually rising pattern was linked to more favorable results. Based on this observation, the five original classes were consolidated into three broader patterns: low-declining (Group 1), moderate-rising (Group 2), and persistently high (Group 3)**(Figure 4)**. Compared with the low-declining group, the moderate-rising group showed a significantly lower risk of unfavorable outcomes (OR = 0.42, 95% CI 0.23-0.78; P = 0.006), whereas the persistently high group did not differ significantly (OR = 0.86, 95% CI 0.48-1.67; P = 0.648). These findings suggest that patients whose SBP remains above 140 mmHg with a gradual upward trend tend to achieve better functional outcomes than those whose SBP stays below 140 mmHg and continues to decline**(Table S6)**.

**Figure 4:**
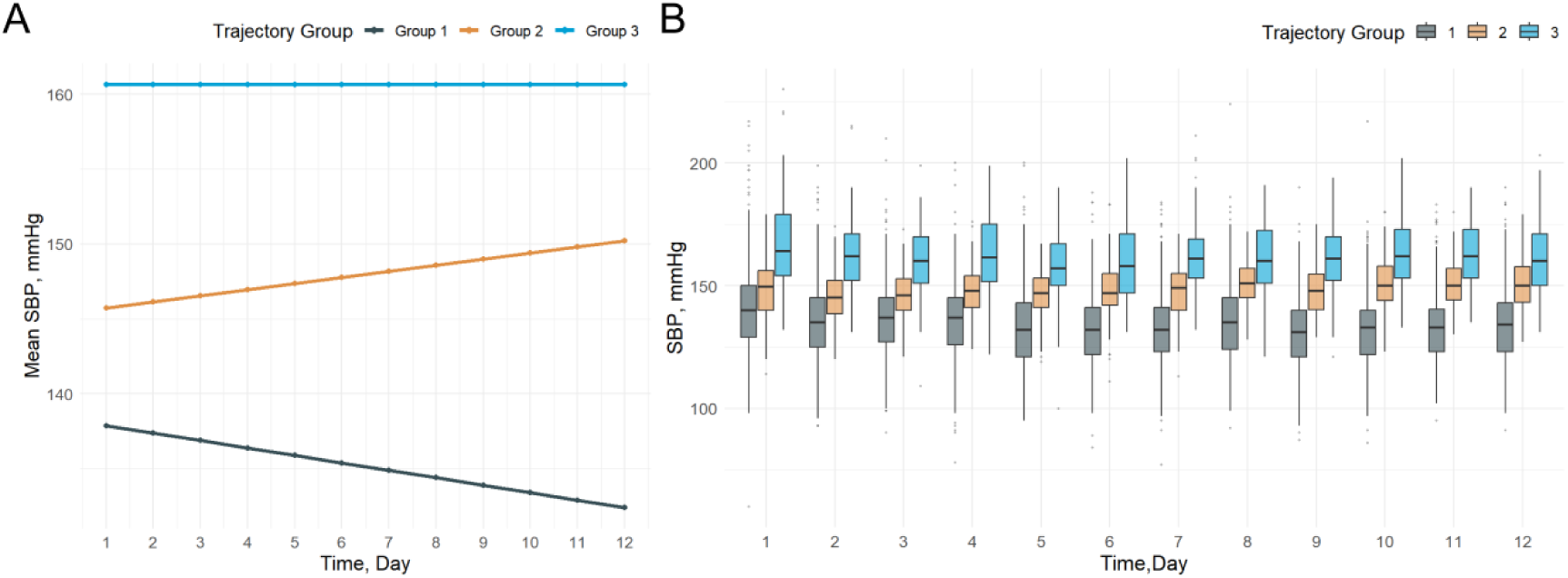
Trajectories of Systolic blood pressure (SBP) during the first 3 days after operation. (A) Model-based estimated SBP trajectories over 12 time points, depicting three distinct trajectory groups: low-declining (Group 1), moderate-rising (Group 2), and persistently high (Group 3). (B) Observed SBP distributions at each time point stratified by the three trajectory groups.

## DISCUSSION

In this study, using continuous blood pressure monitoring during the first 3 postoperative days, we systematically evaluated the associations of early postoperative blood pressure levels, variability, and longitudinal trajectories with functional outcomes in postoperative hypertensive patients with aSAH. Several interesting observations were noted. First, lower SBP during the first 3 days after surgery were strongly associated with an increased risk of unfavorable functional outcome. Second, maintaining SBP ≥ 140 mmHg was associated with better outcomes, about 35% of which was mediated by reduced DCI, indicating potential harm from excessive BP reduction. Third, higher BP variability (SD and CV) independently predicted worse outcomes. Finally, SBP trajectories showed that levels maintained above 140 mmHg with a gradual rise were linked to better outcomes than persistently low and declining SBP.

Blood pressure management plays a critical role in the functional recovery of patients with aSAH5–8. However, most studies address preoperative BP control25, 26, with limited and low-quality evidence for the postoperative phase, especially in patients with postoperative hypertension. The HIMALAIA trial, one of the few randomized controlled studies evaluating induced hypertension for delayed cerebral ischemia after aneurysm repair, was terminated prematurely because it failed to demonstrate any improvement in long-term functional outcomes and raised safety concerns due to a higher incidence of serious adverse events; notably, the trial did not specifically target patients with postoperative hypertension13, 27.In this study, we specifically examined postoperative hypertensive patients and systematically evaluated the relationship between BP levels within the first 3 days after surgery and functional outcomes. We found that lower minimum SBP during the first 3 days were strongly associated with unfavorable outcomes, highlighting the importance of maintaining adequate perfusion pressure during this critical window. Furthermore, daily monitoring of the lowest SBP revealed that patients with a progressive decline in blood pressure over the first three postoperative days experienced significantly worse outcomes, suggesting that excessive and sustained blood pressure reduction may be harmful in this population. Notably, maintaining postoperative SBP≥140 mmHg was linked to lower risks of unfavorable outcomes, with about 35% of the benefit mediated through DCI reduction. These findings suggest that excessive postoperative BP lowering in hypertensive aSAH patients may compromise cerebral perfusion, increase the risk of DCI, and ultimately worsen recovery. In summary, our findings are consistent with previous studies, underscoring that maintaining an appropriate blood pressure level in the postoperative period is crucial for preserving cerebral perfusion and improving outcomes27. This association can be partly explained by the unique pathophysiological alterations following aSAH28. Clinical studies have shown that early impairment of autoregulation substantially increases the risk of cerebral vasospasm and DCI 29, 30. Experimental data further indicate that SAH markedly narrows the autoregulatory range and reduces baseline cerebral blood flow by over 30%, rendering the brain increasingly dependent on blood pressure to maintain perfusion even within previously acceptable ranges31. Under these conditions, maintaining a moderately elevated postoperative blood pressure may help sustain cerebral perfusion and mitigate the risk of DCI. Consistent with this concept, recent studies have indicated that BP management guided toward the patient-specific optimal cerebral perfusion pressure, which reflects the range where cerebrovascular autoregulation is most effective, can improve cerebral blood flow and is associated with better neurological outcomes in patients vulnerable to DCI.32, 33.

Blood pressure variability has an important impact on patient outcomes. In this study, higher SD and CV of SBP were independently associated with unfavorable outcomes. This finding is consistent with previous reports. Xu et al. demonstrated that greater postoperative sBP variability during the first 24 hours after coil embolization predicted poor functional status at discharge, irrespective of mean BP levels34. Besides, Cai et al. found that, among poor-grade aSAH patients, smaller BP fluctuations were associated with better neurological recovery, whereas higher variability markedly increased 6 month mortality35. Moreover, a large cohort study also found that increased BP variability was closely associated with higher in-hospital mortality and unfavorable discharge disposition8. Such fluctuations may also impair cerebral perfusion, thereby providing a plausible explanation for the strong association between postoperative BP variability and poor outcomes.

Finally, this study used trajectory modeling to analyze postoperative blood pressure dynamics and demonstrated their prognostic value. Trajectory modeling, a statistical method that identifies latent subgroups from longitudinal data, has been applied in cardiovascular disease, hemorrhagic stroke, and intracerebral hemorrhage, but has not previously been used in the context of postoperative BP management for aSAH36–38. Unlike analyses based solely on single time points or mean values, this approach captures temporal patterns of BP change, providing insights more closely aligned with the underlying disease process. In our study, patients with SBP maintained around 140 mmHg and showing a slow upward trend (moderate-rising) had better outcomes than those with SBP below 140 mmHg and a persistent decline (low-declining). This finding further supports our earlier analysis that maintaining SBP slightly above 140 mmHg with a stable upward trend in the early postoperative period may be more beneficial for cerebral perfusion and functional recovery than persistent low BP. Besides, persistently high SBP showed no prognostic advantage compared with the low-declining group, likely because the potential benefit of increased perfusion was offset by the risk of rebleeding or cardiopulmonary complications13.

There are several limitations to this study. Although 702 patients were included, the study was from a single center and retrospective in nature, so some degree of selection bias is still possible. Blood pressure values came from routine clinical monitoring rather than a uniform research protocol, and the effects of treatment decisions could not be fully accounted for. Finally, as an observational study, the results show associations rather than causation, and these findings will need confirmation in future multicenter prospective studies and randomized trials.

## CONCLUSIONS

In this study of hypertensive patients with aSAH, we found that lower postoperative SBP was strongly associated with unfavorable outcomes, whereas maintaining SBP ≥140 mmHg conferred a clear protective effect, with part of the benefit mediated through reduced DCI. Trajectory analysis reinforced this finding by showing that patients with moderately rising SBP patterns (≥140 mmHg) achieved the most favorable recovery. In addition, greater blood pressure variability was independently linked to worse outcomes. Together, these findings underscore the clinical importance of maintaining adequately elevated and stable postoperative blood pressure to optimize functional outcomes in this high risk population.

## Data Availability

To protect patient privacy, the data from this study cannot be shared at this time.

## Non-standard Abbreviations and Acronyms

aSAH: Aneurysmal subarachnoid hemorrhage
BP: Blood pressure
SBP: Systolic blood pressure
DCI: Delayed cerebral ischemia
MAP: Mean arterial pressure
BPV: Blood pressure variability
SD: Standard deviation
CV: Coefficient of variation
ARV: Average real variability
SV: Successive variation
BIC: Bayesian Information Criterion

## Acknowledgment

None.

